# Locations of burr holes are associated with recurrence after single burr hole drainage surgery for chronic subdural hematoma

**DOI:** 10.1101/2022.01.03.22268707

**Authors:** Hiroaki Hashimoto, Tomoyuki Maruo, Yuki Kimoto, Masami Nakamura, Takahiro Fujinaga, Yukitaka Ushio

**Author notes:** Corresponding Author Hiroaki Hashimoto, Neurosurgeon/Medical staff, Department of Neurosurgery, Otemae Hospital, Osaka, 540-0008, Japan, Corresponding Author’s, <u>; </u>.

## Abstract

**Objective:** This study aimed to reveal the relation between chronic subdural hematomas (CSDH) recurrence and locations of CSDH and burr holes.

**Methods:** Initial single burr hole surgeries for CSDH with a drainage tube between April 2005 and October 2021 at Otemae Hospital were enrolled. Patients’ medical records, CSDH volume, and CSDH computed tomography values (CTV) were evaluated. The locations of CSDH and burr holes were assessed using Montreal Neurological Institute coordinates.

**Results:** We enrolled 223 patients (bilateral CSDH in 34 patients), and 257 surgeries were investigated. Rate of CSDH recurrence requiring reoperation (RrR) was 13.5%. RrR rate was significantly higher in patients aged ≥76 years, bilateral CSDH, and postoperative hemiplegia. In RrR, preoperative CSDH volume was significantly larger, and CTV was significantly smaller. Locations of CSDH had no influence on recurrence. However, in RrR, locations of burr holes were more lateral and more ventral. Multivariate Cox proportional hazards regression analysis showed that bilateral CSDH, more ventral burr hole positions, and postoperative hemiplegia were risk factors for recurrence.

**Conclusions:** Locations of burr holes related to recurrence. In RrR, CSDH profiles had larger volume and less CTV. Hemiplegia after burr hole surgery is a warning sign for RrR.

## Introduction

Neurosurgeons frequently encounter chronic subdural hematomas (CSDHs), which occur in approximately 80–120 per 100,000 persons in the aged population (1, 2) and are predicted to become the most frequent cranial neurosurgical condition among adults in the future (1, 3). Burr hole surgery with a closed drainage system is a simple operation that is effective for CSDH (4-10). However, neurosurgeons sometimes observe CSDH recurrence. Various factors such as age (11, 12), and diabetes mellitus (11) have been reported to be risk factors for recurrence, and recent studies evaluating CSDH quantitatively have revealed that a large volume of preoperative CSDH was a risk factor for recurrence (13, 14). Although previous studies involving qualitative assessments of CSDH have reported that hyperdense CSDH (12, 15, 16) or cranial base CSDH (17) were associated with a risk of recurrence, the quantitative assessment of CSDH density and its location remains to be performed. Furthermore, the quantitative analysis of burr hole locations is limited. We hypothesized that quantitative assessment of density and locations would enable us to reveal a novel factor for CSDH recurrence.

## Methods and materials

### Patients and study setting

In this retrospective study, we enrolled patients who underwent surgical evacuation of CSDHs at Otemae Hospital between April 2005 and October 2021. We only assessed initial surgeries and excluded surgeries for recurrence cases. All cases in this study were diagnosed as CSDH and not hygroma by at least one neurosurgery specialist certified by the Japan Neurosurgical Society. This study was approved by the Ethics Committee of the Otemae Hospital (Osaka, Japan; approval no. CT190122034) and was conducted in accordance with the Declaration of Helsinki for experiments involving humans. Due to the retrospective and noninvasive nature of the study, informed consent was obtained using the opt-out method from our center’s website.

### Surgical procedure

Surgical evacuation was performed for CSDH, which compressed the brain and induced neurological deficits or medically intractable headaches. Under local anesthesia, a single burr hole was performed, and the dura mater was cut. The outer membrane of the CSDH was exposed, and a silicone tube with a closed drainage system was inserted into the subdural space. Irrigation of the hematoma with normal saline and the direction (anterior or posterior) of insertion of the drainage tube were left to the surgeon’s discretion. A postoperative computed tomography (CT) scan was routinely performed on the morning after the operation, and the tube was removed within 48 h following surgery. Until the removal of the tube, we instructed the patients to remain in the supine position in bed.

### Anticoagulant/antiplatelet medications

Postoperative anticoagulant and antiplatelet medications were temporarily discontinued in patients with a low risk of thrombosis. The medications were resumed once active bleeding was ruled out. The study had no strict criteria for the discontinuation and resumption of anticoagulants or antiplatelets. The decision was left to the surgeon’s discretion.

### Data collection

We collected scans from CT which was performed preoperatively, and we carefully confirmed that the diagnosis was CSDH, not a subdural hygroma. We defined the density of a subdural hygroma as the same as that of cerebrospinal fluid, and we visually differentiated CSDH from a hygroma. We did not set an exact limit using Hounsfield units for discrimination between CSDH and hygroma. We retrospectively evaluated medical variables related to patients. We defined symptoms that were observed until one week after the operation as postoperative symptoms. CSDH were classified into four subtypes: homogeneous, laminar, separated, and trabecular, as described by Nakaguchi et al (17). Data related to treatment options, including drainage tube direction (anterior or posterior) and combination with irrigation, were collected. Details were described in Supplemental Methods. Deterioration based on CT imaging (i.e., increase in hematoma volume) and clinical symptoms (i.e., worsening neurological deficits) necessitated a second operation, defined as recurrence requiring reoperations (RrR). The duration from the first surgery to RrR was recorded.

### Quantitatively assessment for CSDH

Using Digital Imaging and Communications in Medicine (DICOM) CT images, CSDH and postoperative air were segmented, and their volume were calculated by MATLAB R2020b (MathWorks, Natick, MA, USA) (Fig.1a and 1b). The averaged CT values (CTVs) and the standard deviation (SD) were calculated from preoperative CSDH segmentations. The drainage ratio of CSDH was calculated. We defined the thickness (mm) of preoperative CSDH as the maximum horizontal width of preoperative CSDH segmentations (Fig.1c). The homogeneous subtypes were divided into three types according to the average CTV: hypodensity < 30, 30 ≤ isodensity < 40, and 40 ≤ hyperdensity.

**Fig. 1.**
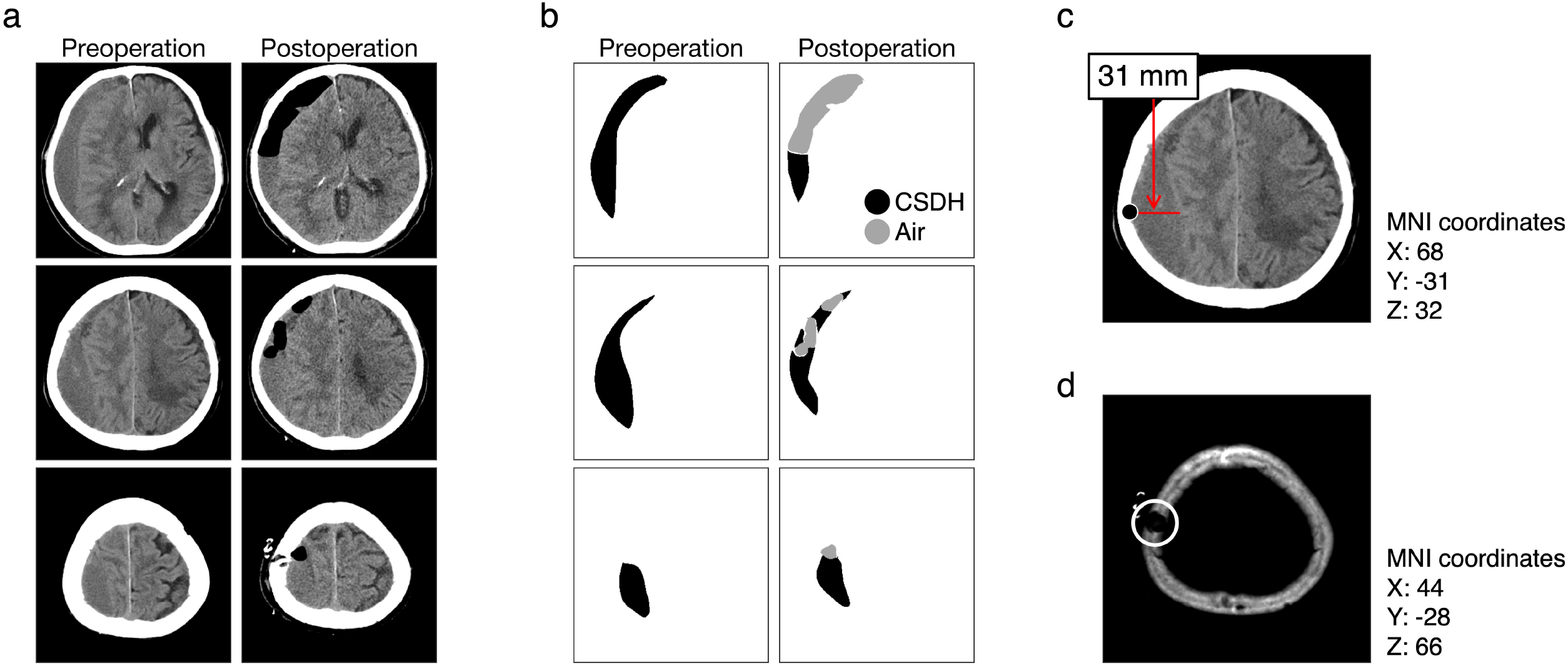
Processing of DICOM CT images of patients with CSDH **a**. CT images of a same CSDH case preoperatively, and postoperatively. **b**. Segmentation of CSDH and air are indicated by black and gray colors, respectively. The shown segmentations were acquired from the images in the panel a. **c**. The maximum width of CSDH is indicated by a red line, which is 31 mm. We treated this width as the thickness of CSDH. The most lateral position is indicated as a black-filled circle, whose MNI coordinates are shown. This image is the same as the middle preoperative image in the panel a. **d**. The position of a single burr hole is indicated as a white circle, whose MNI coordinates are shown.

DICOM CT images were converted into the standard brain using the Montreal Neurological Institute (MNI) coordinates using Brainstorm software (http://neuroimage.usc.edu/brainstorm/). The MNI coordinates for the most lateral position of the CSDH thickness (indicated by a black dot in Fig.1c) and burr hole (indicated by a white circle in Fig.1d) were collected. We treated the thickness positions as a representation of the CSDH locations. We changed the X coordinates to absolute values for the evaluation. The distance between the thickness of the CSDH and the burr hole was calculated. Details were described in Supplemental Methods.

### Statistical analyses

Categorical data and continuous variables were presented as frequencies (percentages) or mean ± SD, respectively. Clinical differences between non-RrR (nRrR) and RrR were assessed using the chi-squared test for categorical variables. Since the distribution of continuous variables was a non-parametric distribution, we used the two-tailed Wilcoxon rank-sum test for continuous variables. The Kruskal-Wallis test was used to compare the four subtype groups. We calculated the Spearman correlation coefficients between the continuous variables.

The receiver operating characteristic (ROC) curve was used to evaluate predictors of RrR. The cut-off value was defined as the maximal Youden index (sensitivity + specificity - 1). Kaplan-Meier survival curves were plotted, and the log-rank test was conducted. Bonferroni correction was used for solution of multiple comparisons. Univariate and multivariable Cox proportional hazards regression analyses were used to calculate hazard ratios (HRs) with 95% confidence intervals (CIs) for RrR. Details were described in Supplemental Methods.

### Bilateral CSDH

Bilateral CSDH surgeries were counted as two different surgeries. In bilateral CSDH onset, almost patients underwent bilateral burr hole surgeries simultaneously; in one patient with bilateral CSDH onset, only one side was treated (Supplemental Fig.1a). The factors that were one value per patient, such as age, sex, and symptoms, were statistically handled based on patients (Supplemental Fig.1b). However, the factors that were one value per hematoma, such as CSDH volume and CT values, were statistically handled based on surgeries (Supplemental Fig.1c). In Cox proportional hazards regression analyses, we treated bilateral CSDH cases based on patients. If the right and left hematoma of bilateral CSDH showed simultaneous recurrence or no recurrence, we used averaged values of CSDH volume, CTV, and MNI coordinates for Cox proportional hazards regression analyses. If bilateral CSDH showed hemilateral recurrence, we used values obtained from the recurrence side (Supplemental Fig.2).

### Data availability

All data in this study are available from the corresponding authors upon reasonable request and after additional ethics approval.

## Results

### Baseline characteristics

Data on a total of 257 surgeries performed in 223 patients were collected (Supplemental Fig.1a). In 37 cases, postoperative clinical information was not available, and in 38 cases, preoperative clinical information was not available (Supplemental Fig.1b). Otemae Hospital did not have an electronic medical records system prior to 2007, and paper-based medical records from 2005 until 2007 could not be obtained.

Of the 223 patients (mean age 76.9 years; range 49–95 years; 59 females, 26.5%), 30 patients (13.5%) experienced RrR, and the median interval between initial surgery and RrR was 31 days. All cases underwent single burr hole surgery. No patients have undergone other surgical operations for CSDH, such as twist drill, two burr holes, craniotomy, and middle meningeal artery embolization, in Otemae Hospital.

Descriptive results based on patients are shown in Table 1. Over 76 years old, bilateral CSDH, and postoperative hemiplegia were significantly associated with RrR. A prescription of anticoagulant and/or antiplatelet medication and preoperative symptoms did not influence RrR.

**Table. 1.** Descriptive statistics regarding demographic characteristics, and pre- and postoperative clinical information calculated from 223 patients. *P* values were calculated using the chi-squared test for categorical variables, and the Wilcoxon rank-sum test for continuous variables. Statistically significant *p* values are flagged with an asterisk.

| Characteristics analyzed | Patients n<br>(%) | nRrR<br>(%) | RrR<br>(%) | *p*-value |
| --- | --- | --- | --- | --- |
| **Sex** |  |  |  |  |
| Male | 164 (74) | 141 (86) | 23 (14) | 0.68 |
| Female | 59 (26) | 52 (88) | 7 (12) |  |
| **Mean age ± SD (years)** |  |  |  |  |
| Male | 75.8 ± 10.4 | 75.2 ± 10.4 | 79.0 ± 9.4 | 0.10 |
| Female | 79.9 ± 8.3 | 79.2 ± 8.5 | 84.6 ± 5.0 | 0.10 |
| **Age categories (years)** |  |  |  |  |
| $\leq 76$ | 85 (38) | 79 (93) | 6 (7) | **\*0.028** |
| $> 76$ | 138 (62) | 114 (83) | 24 (17) | |
| **Bilateral CSDH** |  |  |  |  |
| Yes | 35 (16) | 26 (74) | 9 (26) | **\*0.021** |
| No | 188 (84) | 167 (89) | 21 (11) |  |
| **Laterality (188 hemilateral onset)** |  |  |  |  |
| Left | 98 (52) | 86 (88) | 12 (12) | 0.63 |
| Right | 90 (48) | 81 (90) | 9 (10) |  |
| **Midline shift** |  |  |  |  |
| Yes | 173 (78) | 152 (88) | 21 (12) | 0.29 |
| No | 50 (22) | 41 (82) | 9 (18) |  |
| **Career of surgeons (years)** | | 9.7 ± 4.5 | 9.9 ± 4.9 | 0.83 |
| **Clinical information before** |  |  |  |  |
| **operation #1** |  |  |  |  |
| **Headache** |  |  |  |  |
| Yes | 40 (22) | 32 (8) | 8 (20) | 0.46 |
| No | 145 (78) | 123 (85) | 22 (15) |  |
| **Dementia** |  |  |  |  |
| Yes | 39 (21) | 31 (79) | 8 (21) | 0.41 |
| No | 146 (79) | 124 (85) | 22 (15) |  |
| **Aphasia** |  |  |  |  |
| Yes | 9 (5) | 7 (78) | 2 (22) | 0.62 |
| No | 176 (95) | 148 (84) | 28 (16) |  |
| **Hemiplegia** |  |  |  |  |
| Yes | 126 (68) | 107 (85) | 19 (15) | 0.54 |
| No | 59 (32) | 48 (81) | 11 (19) |  |
| **Gait disturbance** |  |  |  |  |
| Yes | 159 (86) | 133 (84) | 26 (16) | 0.90 |
| No | 26 (14) | 22 (85) | 4 (15) |  |
| **GCS** |  |  |  |  |
| 15 | 107 (58) | 94 (88) | 13 (12) | 0.079 |
| 3 – 14 | 78 (42) | 61 (78) | 17 (22) |  |
| **Anticoagulant** |  |  |  |  |
| Yes | 24 (13) | 22 (92) | 2 (8) | 0.26 |
| No | 161 (87) | 133 (83) | 28 (17) |  |
| **Antiplatelet** |  |  |  |  |
| Yes | 37 (20) | 32 (86) | 5 (14) | 0.62 |
| No | 148 (80) | 123 (83) | 25 (17) |  |
| **Both anticoagulant and antiplatelet** |  |  |  |  |
| Yes | 6 (3) | 6 (100) | 0 (0) | 0.27 |
| No | 179 (97) | 149 (83) | 30 (17) |  |
| **Either anticoagulant or antiplatelet** |  |  |  |  |
| Yes | 55 (30) | 48 (87) | 7 (13) | 0.40 |
| No | 130 (70) | 107 (82) | 23 (18) |  |
| **Clinical information after operation \*2** |  |  |  |  |
| **Complication** |  |  |  |  |
| Yes | 12 (6) | 9 (75) | 3 (25) | 0.39 |
| No | 174 (94) | 147 (84) | 27 (16) |  |
| **Dementia** |  |  |  |  |
| Yes | 28 (15) | 22 (79) | 6 (21) | 0.41 |
| No | 158 (85) | 134 (85) | 24 (15) |  |
| **Hemiplegia** |  |  |  |  |
| Yes | 5 (3) | 0 (0) | 5 (100) | $*2.4 \times 10^{-7}$ |
| No | 181 (97) | 156 (86) | 25 (14) |  |
| **Gait disturbance** |  |  |  |  |
| Yes | 19 (10) | 13 (68) | 6 (32) | 0.053 |
| No | 167 (90) | 143 (86) | 24 (14) |  |
| **GCS score** |  |  |  |  |
| 15 | 150 (81) | 128 (85) | 22 (15) | 0.27 |
| 3–14 | 36 (19) | 28 (78) | 8 (22) |  |

Abbreviations: CSDH, chronic subdural hematoma; GCS, Glasgow Coma Scale; nRrR, non-recurrence requiring reoperation; RrR, recurrence requiring reoperation; SD, standard deviation.
\*1: 185 patients were analyzed. \*2: 186 patients were analyzed. These are indicated in Supplemental Fig.1b.

### Quantitative measurement

Descriptive results based on surgeries are shown in Table 2. Preoperative, and postoperative CSDH volume of RrR was significantly larger than that of nRrR (*p* = 0.024, and 0.00036, respectively; Wilcoxon rank-sum test). Contrarily, the CTV of preoperative CSDH and the drainage ratio were significantly lower in RrR than in nRrR cases (*p* = 0.010, and 0.027, respectively; Wilcoxon rank-sum test). Preoperative CSDH thickness, SD values of CTV, and postoperative air volume showed no significant differences. Preoperative CSDH volume was positively correlated with postoperative CSDH volume, CSDH thickness, age, air volume, and CTV (Supplemental Fig.3a). The regression line between preoperative CSDH volume and thickness was defined by the following equation: preoperative CSDH volume (ml) = 4.3 × thickness (mm) −1.2 (Supplemental Fig.3b).

**Table 2.** Descriptive statistics regarding operative information, and quantitative measures using DICOM CT images calculated from 257 surgeries. *P* values were calculated by the chi-squared test for categorical variables, the Wilcoxon rank-sum test for continuous variables, and the Kruskal-Wallis test for the four subtype groups. Statistically significant *p* values are flagged with an asterisk.

| Characteristics analyzed | Patients n<br>(%) | nRrR<br>(%) | RrR<br>(%) | *p*-value |
| --- | --- | --- | --- | --- |
| **Preoperative CSDH volume ± SD (ml)** | | 116.1 ± 36.6 | 134.0 ± 38.8 | **\*0.024** |
| **Preoperative CSDH thickness ± SD (mm)** | | 28.0 ± 5.1 | 28.6 ± 6.9 | 0.90 |
| **Preoperative CTV ± SD** | | 34.4 ± 6.9 | 31.1 ± 7.7 | **\*0.010** |
| **Preoperative SD of CTV ± SD** | | 9.6 ± 2.6 | 10.3 ± 2.7 | 0.12 |
| **Subtype** |  |  |  |  |
| Homogeneous | 104 (40) | 89 (86) | 15 (14) | 0.053 |
| Laminar | 16 (6) | 13 (81) | 3 (19) |  |
| Separate | 64 (25) | 52 (81) | 12 (19) |  |
| Trabecular | 73 (28) | 70 (96) | 3 (4) |  |
| **Subtype CTV ± SD** |  |  |  |  |
| Homogeneous | 34.8 ± 8.9 | | | **\*0.024** |
| Laminar | 35.6 ± 5.9 | | | |
| Separate | 33.7 ± 5.6 | | | |
| Trabecular | 32.8 ± 5.1 | | | |
| **SD of subtype CTV ± SD** |  |  |  |  |
| Homogeneous | 8.2 ± 1.9 | | | **\*1.3 $\times 10^{-17}$** |
| Laminar | 9.6 ± 2.6 | | | |
| Separate | 11.9 ± 2.4 | | | |
| Trabecular | 9.9 ± 2.2 | | | |
| **Sub-grading in homogeneous group \*1** |  |  |  |  |
| Hypodensity (CTV < 30) | 25 (24) | 18 (72) | 7 (28) | 0.082 |
| Isodensity ($30 \leq \text{CTV} < 40$) | 45 (43) | 40 (89) | 5 (11) | |
| Hyperdensity (CTV $\geq 40$) | 34 (33) | 31 (91) | 3 (9) | |
| **MNI coordinates of thickness ± SD** |  |  |  |  |
| X (absolute values) | | $53.4 \pm 9.9$ | $51.8 \pm 11.5$ | 0.52 |
| Y | | $1.8 \pm 39.8$ | $12.2 \pm 39.4$ | 0.16 |
| Z | | $29.5 \pm 18.2$ | $28.5 \pm 16.5$ | 0.60 |
| **MNI coordinate of burr holes ± SD** |  |  |  |  |
| X (absolute values) | | $55.3 \pm 7.3$ | $58.2 \pm 7.3$ | **\*0.012** |
| Y | | $-5.4 \pm 18.0$ | $-5.6 \pm 22.2$ | 0.62 |
| Z | | $43.6 \pm 8.9$ | $39.2 \pm 11.1$ | **\*0.019** |
| **Distance ± SD** | | $41.4 \pm 24.3$ | $43.3 \pm 27.5$ | 0.93 |
| **Postoperative CSDH volume ± SD (ml)** | | $47.2 \pm 28.2$ | $68.2 \pm 34.1$ | **\*0.00036** |
| **Drainage ratio between pre- and postoperative ± SD** | | $0.59 \pm 0.22$ | $0.48 \pm 0.23$ | **\*0.027** |
| **Postoperative air ± SD (ml)** | | $11.2 \pm 18.6$ | $11.0 \pm 11.7$ | 0.17 |
| **Drainage tube direction** |  |  |  |  |
| Anterior | 110 (43) | 93 (85) | 17 (15) | 0.28 |
| Posterior | 147 (57) | 131 (89) | 16 (11) |  |
| **Irrigation \*2** |  |  |  |  |
| Do | 60 (28) | 52 (87) | 8 (13) | 0.60 |
| Undo | 154 (72) | 129 (84) | 25 (16) |  |

CSDH, chronic subdural hematoma; CTV, computed tomography values; MNI,
Montreal Neurological Institute; nRrR, non-recurrence requiring reoperation; RrR,
recurrence requiring reoperation; SD, standard deviation.
\*1: 104 surgeries classified into the homogenous subtype were analyzed.
\*2: 214 surgeries were analyzed, which are indicated in Supplemental Fig. 1c.

In the four CSDH subtypes, there was no statistical difference in RrR, but the laminar and separate types showed highest recurrence rates (19% and 19%, respectively) and the trabecular type showed lowest recurrence rates (4%). This is concordant with the findings of a previous study (17). There were significant differences in CTV and SD of CTV between the four subtypes (*p* = 0.024, and 1.3 × 10^−17^, respectively; Kruskal-Wallis test). In the homogeneous subtype, the hypodense hematoma was associated with a higher recurrence rate (28%) than the isodense (11%) and hyperdense hematomas (9%), but the difference was not significant.

### Locations of CSDH and burr hole

In the MNI coordinates of the thickness, there were no statistical differences between RrR and nRrR cases (Table 2). All thickness and burr hole positions were plotted over the standard brain (Fig.2). The distribution patterns of thickness positions varied widely in both nRrR and RrR cases. In contrast, the X and Z MNI coordinates of burr hole positions showed significant differences. The X was higher (more lateral), and the Z was lower (more ventral) in RrR (*p* = 0.012, and 0.019, respectively; Wilcoxon rank-sum test) (Fig.2 and Supplemental Fig.4).

**Fig. 2.**
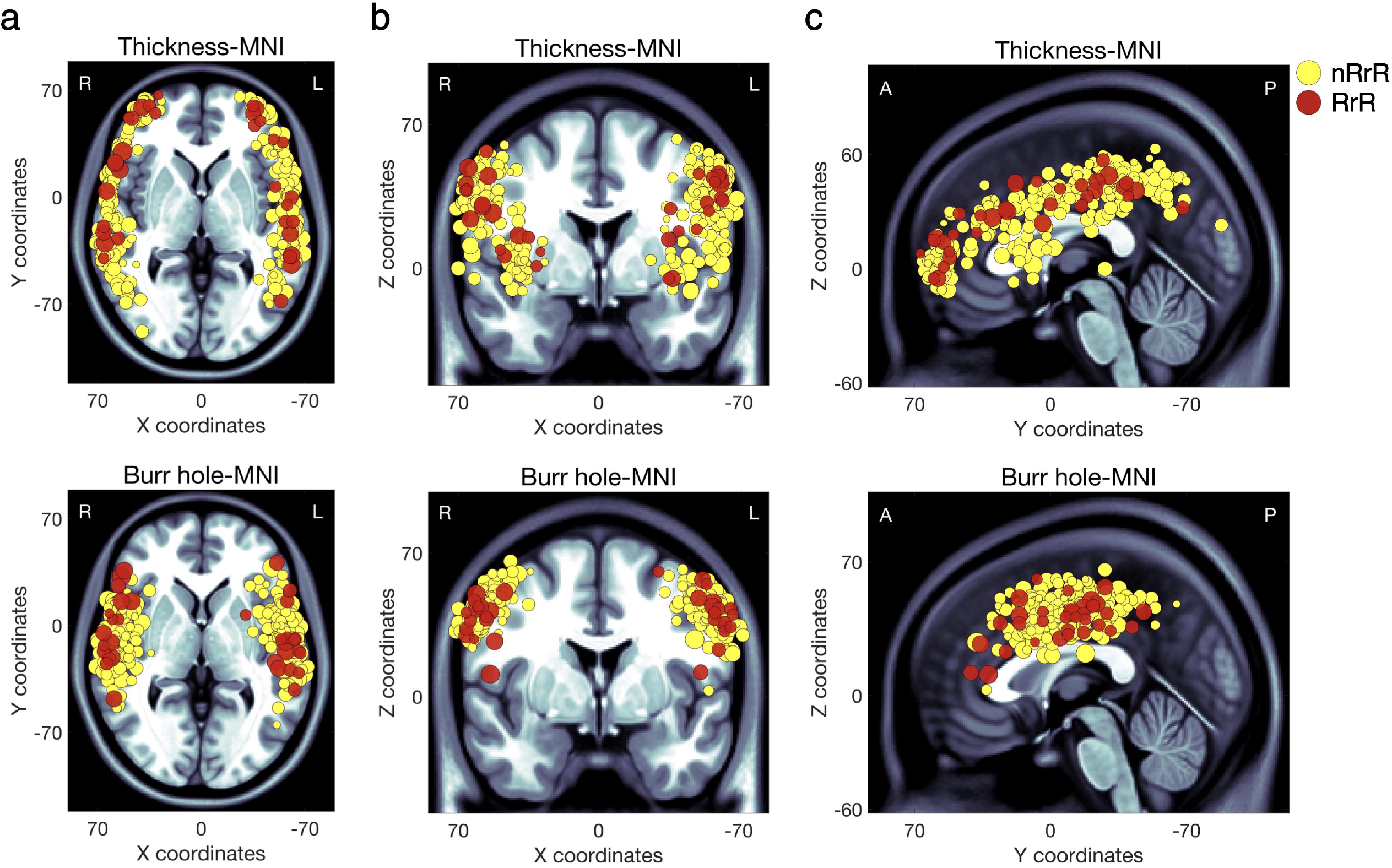
Distribution of CSDH thickness and burr hole positions According to Montreal Neurological Institute (MNI) coordinates, the thickness position of CSDH (upper row) and burr hole position (lower row) are plotted over the MNI standard brain. Cases of recurrence requiring reoperation (RrR) and non-RrR (nRrR) are indicated as red and yellow circles, respectively. The circles are scaled linearly to the preoperative CSDH volume. Axial slices (X-Y plane), coronal slices (X-Z plane), and sagittal slices (Y-Z plane) are indicated in panels a, b, and c. R, right; L, left; A, anterior; P, posterior.

### Follow-up outcomes

According to the Youden index, the ROC analysis showed that the optimal cut-off values were 165 ml for preoperative CSDH volume, 32.5 for CTV, and 76 years for age (Fig.3, upper row). The Kaplan-Meier curve using these cut-off values showed that most RrR cases occurred within approximately 80 days. Log-rank tests revealed that larger preoperative CSDH volume, and lesser CTV were associated with a significant risk of RrR (corrected *p* = 0.0030, and corrected *p* = 0.0099 with Bonferroni correction, respectively; Fig.3, lower row).

**Fig. 3.**
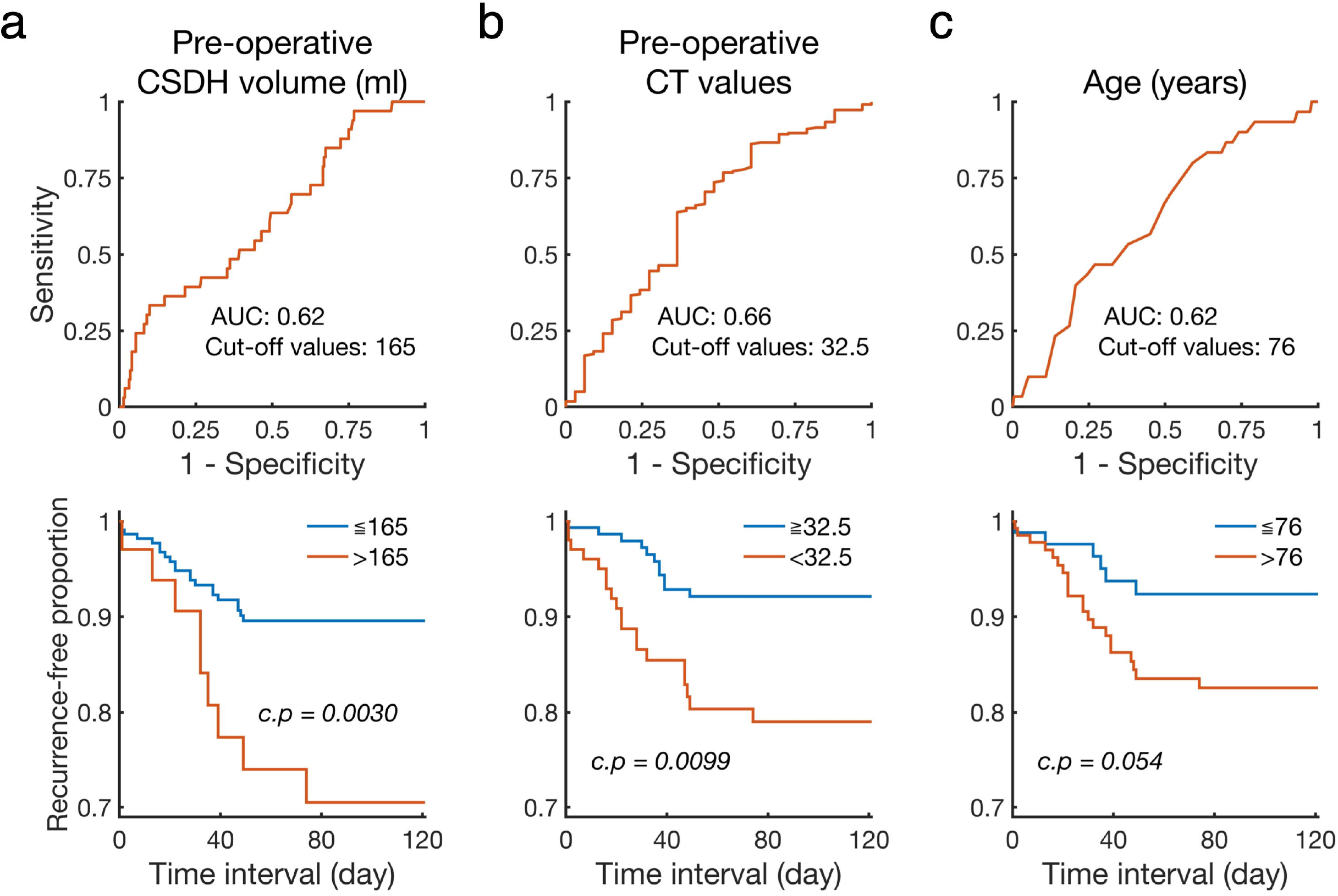
Receiver operating characteristic (ROC) and Kaplan-Meier curves In the upper row, ROC curves are indicated. In the lower row, Kaplan-Meier curves are indicated. Quantitative variables include preoperative CSDH volume (a), CT values (b), and age (c). Area under the curve (AUC) and cut-off values are shown. Kaplan-Meier curves have been plotted using these cut-off values. The results of CSDH volume and CT values were calculated based on 257 surgeries, and the results of age were calculated based on 223 patients. *P* values from the log-rank test are indicated and corrected by Bonferroni correction. c.p, corrected *p* values.

### Risk factors for recurrence of CSDH

Preoperative CSDH volume, age, bilateral CSDH, and postoperative hemiplegia were positively associated with RrR, as revealed by univariate Cox proportional hazards regression analyses. Preoperative CTV and Z coordinates of burr hole positions were inversely associated with RrR. Multivariate Cox proportional hazards regression analyses showed that bilateral CSDH (HR 2.55, *p* = 0.035), Z coordinates of burr hole positions (HR 0.96, *p* = 0.025), and postoperative hemiplegia (HR 17.43, *p* = 0.000012) were independent risk factors for RrR (Table 3).

**Table. 3.** Univariate and multivariate Cox proportional hazards regression analyses of characteristics related to postoperative recurrence requiring reoperation for CSDH. The 186 patients indicated in Supplemental Fig.1b were used. Statistically significant *p* values are flagged with an asterisk.

|  | Cox proportional hazards regression analyses |  |  |  |
| --- | --- | --- | --- | --- |
| Characteristics analyzed | Univariate analysis |  | Multivariate analysis |  |
|  | HR (95% CI) | *p*-value | HR (95% CI) | *p*-value |
| Preoperative CSDH volume (ml) | 1.01 (1.00 - 1.02) | *0.032 | 1.00 (0.99 - 1.01) | 0.59 |
| Preoperative CT values | 0.94 (0.90 - 1.00) | *0.035 | 0.98 (0.92 - 1.04) | 0.45 |
| Age (years) | 1.05 (1.01 - 1.10) | *0.024 | 1.02 (0.98 - 1.07) | 0.33 |
| Bilateral CSDH | 2.45 (1.11 - 5.37) | *0.026 | 2.55 (1.07 - 6.07) | *0.035 |
| X coordinate of burr holes | 1.06 (1.00 - 1.12) | 0.051 | - | - |
| Z coordinate of burr holes | 0.95 (0.92 - 0.99) | *0.0087 | 0.96 (0.92 - 0.99) | *0.025 |
| Postoperative hemiplegia | 17.66 (6.52 - 47.82) | *0.000000<br>016 | 17.43 (4.84 - 62.76) | *0.000012 |

Abbreviations: CSDH, chronic subdural hematoma; CT, computed tomography; CI, confidence interval; HR, hazard ratio

## Discussion

We hypothesized that quantitative assessment of density and locations would enable us to reveal a novel factor for CSDH recurrence. By assessing quantitative values acquired from DICOM CT images of CSDH patients who underwent an initial single burr hole surgery with a closed drainage system, we could show that CTV was significantly lower in RrR cases than in nRrR cases, and a hypodense subgroup (CTV < 30) of homogeneous CSDH cases were associated with a higher RrR rate than iso- and hyperdense subgroups. Regression analyses showed that higher CTVs in CSDHs were associated with a lower risk of RrR. This result is novel, contrary to the results of previous studies indicating that hyperdense hematoma was associated with recurrence (12, 13, 15, 16). We inferred that our results might be related to the poor surgical outcomes associated with hygroma. A subdural hygroma is thought to be a collection of cerebrospinal fluid without blood due to traumatic arachnoid tearing (17, 18), and CT images of hygroma show hypodensity (19, 20). Nearly 50% of patients with a subdural hygroma develop CSDH (18), and the effect of surgery for hygroma is poor (21, 22). Although previous studies assessed CSDH density qualitatively, our study assessed CSDH quantitatively. Moreover, we succeeded in demonstrating statistical differences among four CSDH subtypes using CTV and its SD. These results support the correctness and reliability of our quantitative analyses.

Additionally, we demonstrated that more lateral and more ventral burr hole positions were associated with RrR. Empirically, burr holes were created overlying the hematoma and were mainly on the temporal line, ranging in location from the frontal to the temporal lobe (5). However, there have been no previous studies assessing burr hole positions quantitatively, and the relation between burr hole positions and RrR has remained unknown. According to our result, we propose that making a burr hole located a bit more parietal to the temporal line, avoiding the temporal muscle, would be effective for reducing RrR.

In this study, 13.5% of the participants had RrR, and most CSDHs recurred within 80 days. These findings were concordant with previous studies (11, 12, 16, 23). Several factors reportedly influence recurrence, including age, sex, diabetes mellitus, smoking, bilateral CSDH, midline shift, GCS score at admission, and high-density lipoprotein levels (11, 12, 15, 23-25). We also demonstrated that age and bilateral CSDH influenced recurrence, in agreement with the results of previous studies. Since the career years of operating surgeons was not significantly different between nRrR and RrR cases, we infer that a burr hole surgery for CSDH is a safe procedure for neurosurgical residents to perform. The recurrence rate of cranial base CSDH was reported to be high (17). However, our results demonstrated that the location of CSDH did not influence recurrence. Additionally, we demonstrated that if patients showed postoperative hemiplegia even after burr-hole surgery, this symptom was highly associated with RrR. If hemiplegia continues after the operation, it is reasonable that neurosurgeons try to perform re-operation for relief of the symptom.

Preoperative CSDH volume was a factor for recurrence (12-14). We showed that a large volume of preoperative CSDH was associated with recurrence and the optimal cut-off value for preoperative CSDH volume was 165 ml. However, in a clinical setting, it is difficult to determine CSDH volume easily and rapidly using DICOM CT images, in contrast to the thickness of the CSDH. Therefore, we showed the regression line and calculated the thickness (39 mm) corresponding to 165 mL (Supplemental Fig.3b). Therefore, we propose that physicians monitor for possible recurrence in cases of CSDH with thicknesses of over 39 mm.

Preoperative CSDH volume was positively correlated with several parameters (Supplemental Fig.3a). Postoperative CSDH volume (13, 26), CSDH thickness (6, 15, 27), and postoperative air volume (28, 29) were associated with recurrence. However, the above three parameters were positively correlated with preoperative CSDH volume, and we infer that the true factor influencing recurrence is preoperative CSDH volume.

Although postoperative air volume was previously thought to be a risk factor for recurrence (28, 29), our results showed that postoperative air volume was not a risk factor for recurrence and positively correlated with preoperative CSDH volume. It is reasonable to postulate that the larger the preoperative CSDH volume, the higher the recurrence rates, and the larger the postoperative air volume. Persistence of subdural air after surgery has been significantly correlated with poor brain re-expansion (30), and patients with residual subdural air seven days after surgery have been shown to have a higher recurrence rate than those without air (31). Moreover, poor brain re-expansion seven days after surgery has been shown to be a significant risk factor for recurrence (23), and cerebral atrophy is also associated with the development of CSDH (32). Sufficient subdural space is thought to be a prerequisite for developing CSDH (18). Therefore, we infer that a true risk factor for recurrence of CSDH may be poor re-expansion of the brain, and postoperative air volume may be the result, and not a cause, of poor brain re-expansion.

As for the surgical technique, twist drill craniostomy and double burr hole craniostomy may be performed in the management of CSDH (5). However, we think that these operations are considered minor in Japan, and the single burr hole is the surgery of choice for almost all cases of CSDH. In our institution, all cases of CSDH were treated by single burr hole surgery. Furthermore, the technical aspects of the single burr hole surgeries did not vary during the study period, as confirmed by our operative notes. Some previous studies have proposed that a drainage tube should be inserted into the frontal convexity (31). However, our results showed that the direction of the drainage tube had no influence on recurrence. This result also supports the notion that postoperative air is not a risk factor for recurrence. There have been conflicting reports showing irrigation may correlate to recurrence (8). However, our results showed no statistically significant effectiveness of irrigation.

This study has some limitations. We focused on only single burr hole surgery, not other types of surgery, such as two-burr hole surgeries or twist drill surgery (5). Thus, the results of this study can be applied to only single burr hole surgeries. Second, there may also be a possibility that the surgical procedure was not consistently performed because we collected long-term data spanning 17 years (2005–2021). However, we believe that neurosurgeons in Japan normally treat CSDH through single burr hole and drainage surgery, as opposed to other types of surgery, and the surgical techniques involved in the procedure had already been established in 2005. Therefore, we were not concerned about the biases which may arise due to changes in the surgical procedure over 15 years. Third, due to the retrospective nature of the study, there were no concrete criteria for the use of anticoagulant or antiplatelet medications. This prevents us from precisely assessing the influence of these medications on recurrence. To solve these problems, we think that a prospective study would be needed. Finally, since burr holes were made in the area that neurosurgeons thought that they were effective for drainage, there would be some bias in the results of MNI location of burr holes.

In conclusion, age, bilateral CSDH, preoperative CSDH volume, CTV of the hematoma, and more ventral burr hole positions were related to CSDH recurrence. In cases of low concentration and large volume CSDH, attention should be paid to recurrence.

## Acknowledgments

We would like to thank Dr. Masayuki Hirata at Department of Neurological Diagnosis and Restoration in Graduate School of Medicine, Osaka University for his technical support, Dr. Haruhiko Kishima at Department of Neurosurgery in Graduate School of Medicine, Osaka University for his comment, and Dr. Hiroyuki Kurakami and his colleague at Data Coordinating Center in Osaka University Hospital for their statistical support.

## Authors’ Contributions

HH conceived this study, collected the data, created the MATLAB program, analyzed the data, created all figures, and was primarily responsible for writing the manuscript. HH and TM discussed the interpretation of the results. TM, and YU supervised the study. All authors reviewed the manuscript.

## Supplemental Material

Supplemental Methods

Supplemental Figure 1 – 4.

