## Supplemental Table, Figure 1-4 for "Locations of burr holes are associated with recurrence after single burr hole drainage surgery for chronic subdural hematoma"

Supplemental Methods

Supplemental Figure. 1

Supplemental Figure. 2

Supplemental Figure. 3

Supplemental Figure. 4

**Data collection**

We retrospectively evaluated medical variables related to patients, including sex, age, antiplatelet medication, anticoagulant medication, and pre- and postoperative symptoms such as headache, dementia, aphasia, hemiplegia, gait disturbance, and consciousness level represented by the Glasgow Coma Scale (GCS). Data on hematoma features including hematoma laterality (left or right), bilaterality, presence of midline shift, and subtypes were collected. Since there was no information about irrigation in 43 surgeries, irrigation data were collected from 214 surgeries (Supplemental Fig.1c). Number of years since the surgeon had started neurosurgical residency was also recorded.

**Quantitatively Assessment for CSDH**

Digital imaging and communication in medicine (DICOM) CT images were imported to MATLAB R2020b (MathWorks, Natick, MA, USA), and CSDH and postoperative air were segmented manually using the image segmenter app in MATLAB (<https://www.mathworks.com/help/images/ref/imagesegmenter-app.html>; Fig. 1a and 1b). These procedures enabled us to calculate the volume and average CT values (CTVs) of the segmented regions. We calculated the volume of CSDH preoperatively, and postoperatively, and the volume of postoperative air. The averaged CTVs were calculated from preoperative CSDH segmentations, and the standard deviation (SD) was also calculated. The drainage volume was the difference obtained from the preoperative CSDH volume minus the postoperative CSDH volume. The drainage ratio was calculated by dividing the drainage volume by the preoperative CSDH volume.

The distance between the thickness of the CSDH and the burr hole was calculated using the following equation:

$$Distance =\sqrt[2]{{(x_{thickness} - x_{burr hole})}^{2} +{(y_{thickness} - y_{burr hole})}^{2}+{(z_{thickness} - z_{burr hole})}^{2}}$$

**Statistical analyses**

Kaplan-Meier survival curves were plotted (<https://github.com/dnafinder/kmplot>, Curve Cardillo G, 2008), and the log-rank test was conducted to compare the time to recurrence (<https://github.com/dnafinder/logrank>, Cardillo G. 2008).

Univariate Cox proportional hazards regression analyses were used to calculate hazard ratios (HRs) with 95% confidence intervals (CIs) for RrR. The variables analyzed in the regression analyses were potential predictors, and all of them showed a *p*-value < 0.05, as determined by the chi-squared test or Wilcoxon rank-sum test. Multivariable Cox proportional hazards regression was performed to ensure that the variables were independently predictive of RrR. Statistical analyses were performed using the Statistical and Machine Learning Toolbox of MATLAB R2020b.

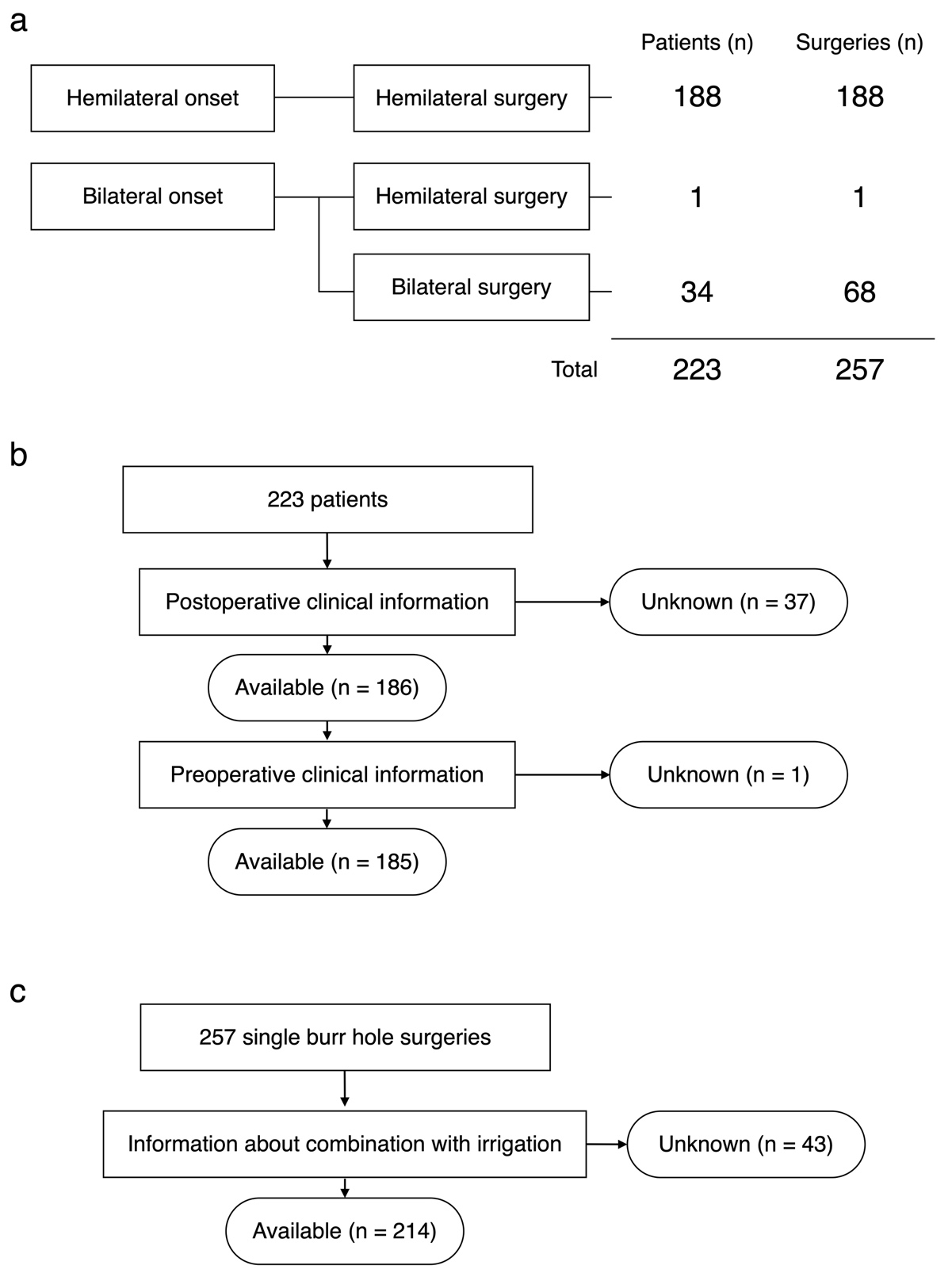
**Supplemental Fig.1** Patients and surgeries investigated in this study.

**a.** We enrolled 223 patients who were suffered from initial onset CSDH including hemilateral onset or bilateral onset. In bilateral onset cases, almost all patients underwent bilateral burr hole surgeries simultaneously. However, one patient underwent a single side burr hole surgery because the CSDH on the other side did not need to be treated. Since simultaneous bilateral burr hole surgeries were treated as two different surgeries, in total 257 surgeries were enrolled. **b.** Flow chart showing 223 patients investigated in this study. **c.** Flow chart showing 257 surgeries investigated in this study.

**Supplemental Fig.2** How bilateral surgery was handled in Cox proportional hazards regression analyses.

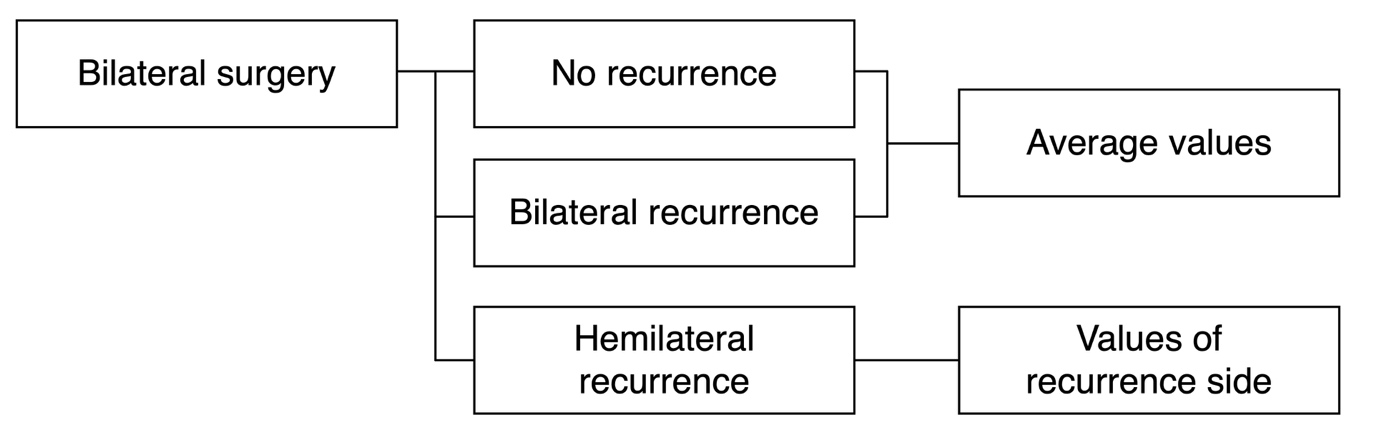

In patients who were underwent simultaneous bilateral surgeries, there were three different outcomes: no recurrence, simultaneous bilateral recurrence, and hemilateral recurrence. In the no recurrence and bilateral recurrence groups, we calculated average values in preoperative CSDH volume, preoperative CT values, and X and Z coordinates of burr holes and used those for Cox proportional hazards regression analyses. In the hemilateral recurrence group, we used values acquired from the recurrence side for Cox proportional hazards regression analyses.

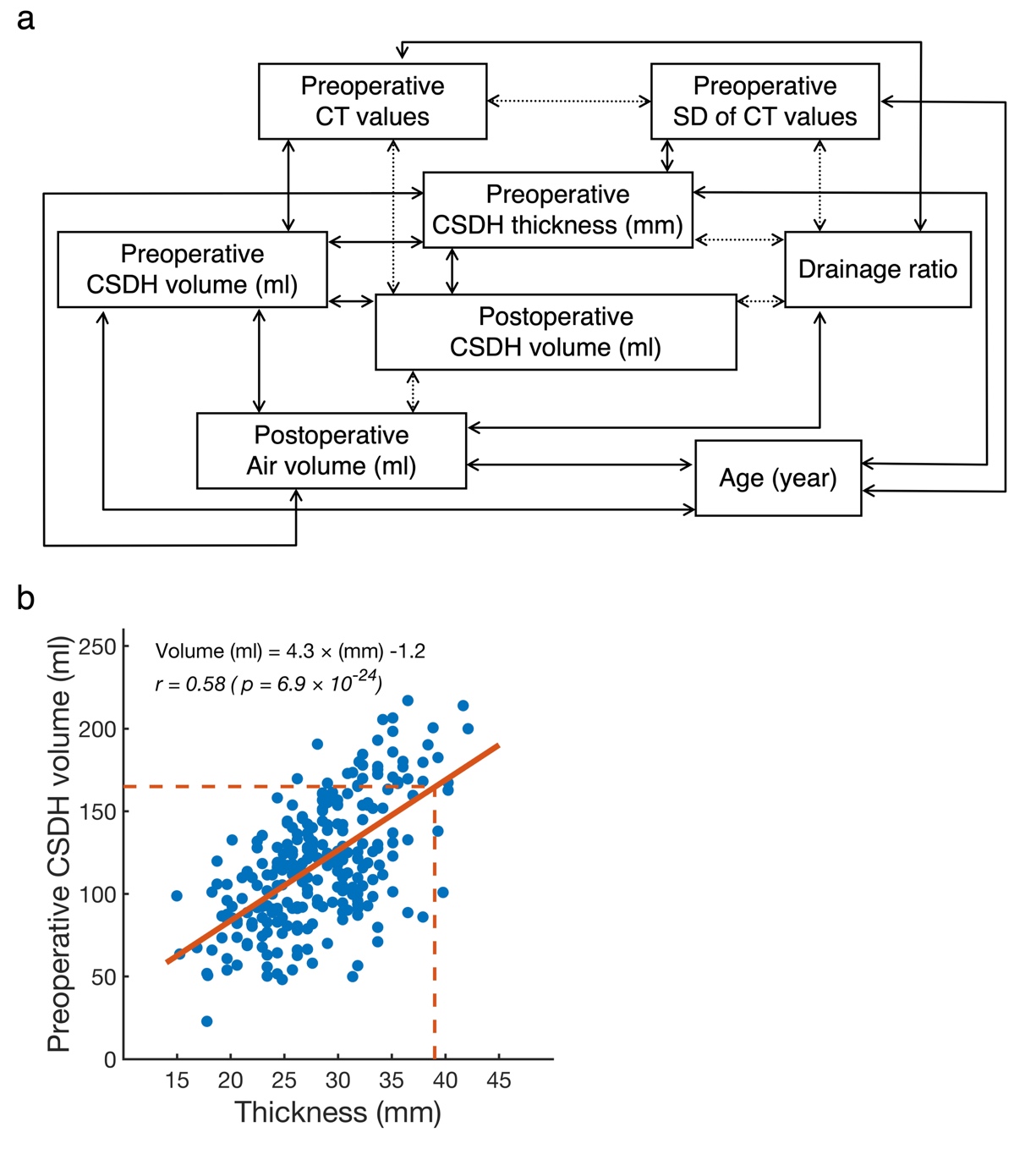
**Supplemental Fig.3** Correlation relations between quantitative variables.

**a.** Correlation relations that indicate *p* values < 0.05 are shown, calculated based on 257 surgeries. Positive and negative correlations are indicated as a solid and a broken line. Since preoperative CSDH volume had a positive correlation with postoperative air volume, we inferred that larger volume CSDH tended to have larger air volume after a burr hole surgery. It was reasonable that the drainage ratio and air volume had a negative correlation with the postoperative CSDH volume. There was a negative correlation between CT values (CTV) and standard deviation (SD) of CTV, which indicated that the higher the hematoma density, the more homogeneous the hematoma density. Postoperative CSDH volume had a negative correlation with CTV, which indicated that the thicker the CSDH concentration, the smaller the postoperative CSDH volume. **b.** Scatter plot between preoperative CSDH volume and thickness are shown. The regression line was indicated as a red line and *r* is the correlation coefficient. Red dashed lines correspond to 39 mm and 165 ml.

**Supplemental Fig.4** Burr holes-related X and Z coordinates indicating significant differences between recurrence requiring reoperation (RrR) and non-RrR (nRrR).

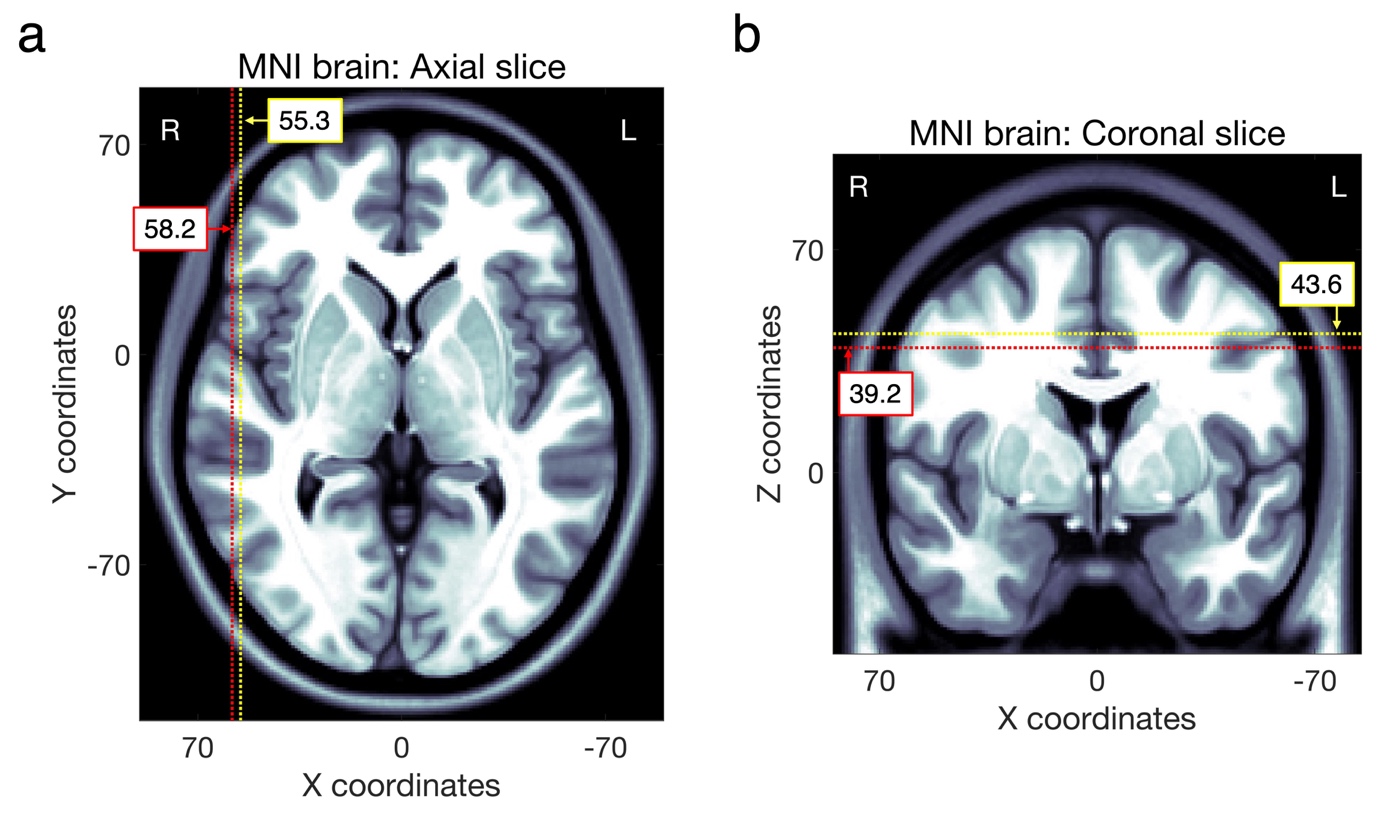

The X and Z burr hole positions indicating significant differences in Table 2 are shown on the Montreal Neurological Institute (MNI) standard brain in the “a” and “b” panels, respectively. The positions related to RrR are indicated by red dashed lines. The positions related to nRrR are indicated by yellow dashed lines.
